# Inpatient Outcomes Of Mechanical Circulatory Support Devices and Bridging to Transplantation in Hypertrophic Cardiomyopathy

**DOI:** 10.1101/2023.06.07.23291114

**Authors:** Bilal Hussain, Constantine Tarabanis, Vishal Dhulipala, Pradeepkumar Devarakonda, Haisum Maqsood, Pedro Moreno

## Abstract

**Objective:** Understand the outcomes of mechanical circulatory support devices (MCSD) and heart transplantation (HT) in patients with underlying hypertrophic cardiomyopathy (HCM).

**Introduction:** HCM can rarely cause severe left ventricular outflow tract obstruction and apical ballooning presenting as cardiogenic shock necessitating the use of mechanical circulatory support devices (MCSD). Data on in-hospital outcomes of HCM patients placed on MCSD and receiving HT is limited.

**Methods:** The National Inpatient Sample (2016-2019) was used for the retrospective analysis of patients hospitalized for MCSD and HT using ICD-10 codes. These patients were divided into two cohorts, with and without HCM, and compared in terms of in-hospital mortality, trends in mortality rates, hospitalization costs and mean length of stay.

**Results:** Among 254170 patients hospitalized for MCSD and HT, 12,000 patients had underlying HCM. Underlying HCM was associated with increased odds of mortality in patients receiving left ventricular assist devices (LVAD) (OR 3.06, 95% CI 1.18-7.93, *p*=0.02) and short-term MCSD (OR 1.8, CI 1.29-2.5, p<0.001. HCM was not associated with increased mortality in patients hospitalized for HT (OR 1.05, CI 0.42-2.6, *p*=0.9). Patients with HCM undergoing MCSD and HT had a longer mean length of stay (26.6 vs 14.4 days, p<0.0001), and higher mean hospitalization charges ($977797 vs $497590, p<0.0001) as compared to non-HCM patients.

**Conclusion:** Underlying HCM is associated with increased in-hospital mortality in patients undergoing LVAD and short-term MCSD placement. Further prospective studies are required to expand our understanding of prognosis of HCM in patients undergoing MCSD and establish management guidelines.

## Introduction

Hypertrophic cardiomyopathy (HCM) is a monogenic cardiac disorder caused by the autosomal dominant inheritance of variants in genes coding cardiac sarcomere proteins or sarcomere-related structures^1^. The pathophysiology of HCM can consist of diastolic dysfunction, dynamic left ventricular outflow tract obstruction (LVOTO), mitral regurgitation (MR) and arrhythmias^2^. Common ensuing complications of HCM include sudden cardiac death (SCD), heart failure (HF) and conduction blocks which are managed with implantation of cardiac devices^2^. Literature suggests that HCM can also rarely present with sudden development of severe and implacable LVOTO in association with severe mitral regurgitation, apical ballooning, decreased left ventricular (LV) ejection fraction, and refractory hypotension leading to cardiogenic shock^2^. Sherrid et al.^3^ described this as ‘Syndrome of Reversible Cardiogenic Shock and Left Ventricular Ballooning in Obstructive Hypertrophic Cardiomyopathy’.

It is crucial to diagnose if HCM is the cause of cardiogenic shock as it is challenging and the features might resemble acute coronary syndrome in an acute presentation. Coronary angiograms will be normal in these patients which might lead to the diagnosis of ‘Takotsubo Cardiomyopathy’ due to apical ballooning, however, these patients will have normal septal thickness, and no LVOTO unlike HCM. Management will be different in both of these patients. Some case studies have been reported for cardiogenic shock in HCM which has been treated with mechanical circulatory support devices (MCSD) including intra-aortic balloon pump (IABP), percutaneous ventricular assist device (Impella), extracorporeal membrane oxygenation (ECMO)^2, 3^. If patient fails these therapies, urgent surgery is also indicated with surgical myomectomy. Relief of obstruction with these measures has shown to improve systolic function and survival^3^. Furthermore, left ventricular assist devices (LVADs) have been described in HCM patients as a bridge to heart transplantation (HT)^4–6^. Data on the impact of MCSD and HT on HCM patient outcomes is limited to case studies^2, 3^. To address this knowledge gap, we focus on data from hospitalizations during which MCSD and HT were performed to characterize the demographic, clinical characteristics and in-hospital outcomes of HCM patients as compared to their non-HCM counterparts.

## Methods

### Data source

Data was obtained from the National Inpatient Sample (NIS), a publicly available deidentified database of hospital inpatient stays in the United States, sponsored by the Agency for Healthcare Research and Quality as part of the Healthcare Cost and Utilization Project. Given the deidentified nature of the data, institutional review board approval was waived. The International Classification of Diseases-10^th^ Edition-Clinical Modification (ICD-10-CM) codes were used to identify all patients aged 18 years or older hospitalized for MCSD and HT from January 1, 2016 to December 31, 2019. MCSD included LVAD and short-term MCSD (IABP, Impella, and ECMO. The study population’s baseline characteristics were obtained using the corresponding ICD-10-CM codes (**Table S1**).

### Study end points

Baseline characteristics and inpatient outcomes were compared between patients with and without HCM who had MCSD and HT performed. Prevalence trends in MCSD and HT in HCM were also investigated, while comorbidity burden was assessed using the Charlson Comorbidity Index (CCI)^7^. The study’s primary outcome was in-hospital mortality compared between the two cohorts. Secondary outcomes included mean length of stay (LOS) and hospitalization costs (adjusted for inflation).

### Statistics

Categorical variables are presented as proportions and compared using Pearson chi-square (X^2^) and Fisher’s exact tests. Continuous variables are presented as mean with standard deviation and compared using Student’ t-test. Univariate logistic regression models were used to investigate the unadjusted effect of underlying HCM on MCSD and HT in-hospital mortality. Multivariable logistic regressions accounted for gender, ethnicity, median household income, insurance, hospital bed-size, teaching status, location, CCI, coronary artery disease (CAD), congestive heart failure (CHF), arrhythmias, chronic pulmonary disease (CPD), liver disease, kidney disease, hypertension (HTN), hyperlipidemia (HLD), and diabetes mellitus (DM). Yearly trends for crude and adjusted mortality rates to account for covariates were tabulated using marginal standardization following logistic regression analyses. The secondary continuous outcomes of LOS and hospitalization costs were compared using multivariable linear regression models adjusting for the same aforementioned variables. All analyses were performed using HCUP recommended stratifying, clustering, and weighting samples with Stata Statistical Software (Version 17, Texas, StataCorp LLC).

## Results

### Baseline characteristics of MCSD and HT patients

Among 254170 patients hospitalized for MCSD, 1290 had underlying HCM. **Table 1** compares the baseline characteristics of patients who had MCSD and HT with and without HCM. Patients who underwent MCSD and HT with underlying HCM had a lower mean age (55 vs. 63.6 years, p<0.001), a higher female proportion (46.5% vs. 29.7%, p<0.001) and a higher proportion with a history of CHF (69.4 vs 56.9%, *p*<0.001) as compared to those without HCM. Hospital characteristics were similar in the two cohorts with hospitalizations predominantly occurring in large urban teaching hospitals in the Southern US. Patients receiving a short-term MCSD had the higher mean age (60.9 years) than those receiving LVAD and HT (42.4, 42 years respectively) **(Table S2).**

**Table 1:** Baseline characteristics for MCSD and HT with and without HCM.

| Patient Characteristic | MCSD and HT Patients (n=254170) |  |  |  | p value |
| --- | --- | --- | --- | --- | --- |
|  | With HCM (n=1290) |  | Without HCM (n=252880) |  |  |
|  | Number of Patients (n) | Proportion (%) | Number of Patients (n) | Proportion (%) |  |
| Age |  |  |  |  |  |
| Mean Age (years, SD) | 55 ±1.4 |  | 63.6 ±0.13 |  | <0.001 |
| Gender |  |  |  |  |  |
| Male | 690 | 53.5 | 193100 | 70.3 | <0.001 |
| Female | 600 | 46.5 | 81615 | 29.7 |  |
| Ethnicity |  |  |  |  |  |
| White | 865 | 69.5 | 185700 | 70.8 | 0.03 |
| African-American | 215 | 17.2 | 30050 | 11.5 |  |
| Hispanic | 80 | 6.4 | 25075 | 9.6 |  |
| Other Ethnicity | 85 | 9.3 | 21650 | 8.1 |  |
| Co-Morbidities |  |  |  |  |  |
| Coronary Artery Disease | 665 | 51.6 | 234905 | 85.5 | <0.001 |
| Congestive Heart Failure | 156305 | 69.4 | 895 | 56.9 | <0.001 |
| Arrhythmia | 990 | 76.7 | 164030 | 59.7 | <0.001 |
| Hypertension | 208755 | 76 | 920 | 71.3 | 0.07 |
| Diabetes | 325 | 40.8 | 112145 | 25.2 | <0.001 |
| Hyperlipidemia | 495 | 38.4 | 151155 | 55 | <0.001 |
| Peripheral Artery Disease | 90 | 7 | 15025 | 5.5 | 0.3 |
| Chronic Pulmonary Disease | 230 | 17.8 | 54635 | 19.9 | 0.4 |
| Liver Disease | 325 | 25.2 | 41375 | 15 | <0.001 |
| Kidney Disease | 153035 | 72.9 | 940 | 55.7 | <0.001 |
| Anemia | 840 | 65 | 137495 | 50 | <0.001 |
| Obesity | 210 | 16.3 | 50930 | 18.5 | 0.3 |
| Smoking | 100 | 7.8 | 51260 | 18.7 | <0.001 |
| Charlson Comorbidity Index |  |  |  |  |  |
| 0 | 65 | 5 | 8930 | 3.3 | 0.001 |
| 1 | 295 | 22.9 | 41900 | 15.3 |  |
| 2 | 290 | 22.5 | 62210 | 22.6 |  |
| ≥3 | 640 | 49.6 | 161700 | 58.9 |  |
| **Median Household Income** |  |  |  |  |  |
| \$1-\$49,999 | 270 | 21.5 | 77310 | 28.7 | 0.054 |
| \$50,000-\$85,999 | 675 | 53.8 | 138035 | 51.3 | |
| >\$86,000 | 310 | 24.7 | 54185 | 20 | |
| **Insurance** |  |  |  |  |  |
| Medicare | 495 | 38.5 | 147580 | 53.8 | <0.001 |
| Medicaid | 185 | 14.4 | 31035 | 11.3 |  |
| Private Insurance | 535 | 41.6 | 75775 | 27.6 |  |
| Others | 70 | 5.5 | 19875 | 7.3 |  |
| **Hospital Characteristics** |  |  |  |  |  |
| **Hospital Bedsize** |  |  |  |  |  |
| Small | 95 | 7.5 | 30155 | 11 | 0.001 |
| Medium | 220 | 17 | 68010 | 24.8 |  |
| Large | 975 | 75.6 | 176575 | 64.2 |  |
| **Location/Teaching Status** |  |  |  |  |  |
| Rural | <11 | 0.8 | 8930 | 3.2 | <0.001 |
| Urban Non-teaching | 95 | 7.4 | 41775 | 15.2 |  |
| Urban Teaching | 1185 | 91.2 | 224035 | 81.5 |  |
| **Region** |  |  |  |  |  |
| Northeast | 265 | 20.5 | 48695 | 17.6 | 0.3 |
| Midwest | 335 | 26 | 64735 | 23.6 |  |
| South | 435 | 33.7 | 109750 | 40 |  |
| West | 255 | 19.8 | 51560 | 18.8 |  |
| **Ownership** |  |  |  |  |  |
| Government | 195 | 15.1 | 26280 | 9.6 | <0.001 |
| Private | 1095 | 84.9 | 248460 | 90.4 |  |

HCM: Hypertrophic Cardiomyopathy, MCS: Mechanical Circulatory Support Devices, HT: Heart
Transplantation

### Prevalence of MCSD and HT in HCM and Trends

Prevalence of MCSD and HT in patients with and without HCM is tabulated in **Figure 1** and **Table 2**. Patients with underlying HCM had consistently higher MCSD and HT rates than those without HCM (LVAD 0.05% vs 0.01%, p<0.001, short-term MCSD 0.4% vs 0.18%, p<0.001) (**Figure 1**). Majority of HCM had short-term MCSD (0.4%), followed by HT (0.2%), and LVAD (0.05%, **Figure 1**). Prevalence of HCM has been steadily trending up from 2016 to 2019 (0.14% to 0.17%, p<0.001) while MCSD and HT rates among HCM patients have remained stable (p>0.05) (**Figure 2, Table S3**).

**Figure 1:**
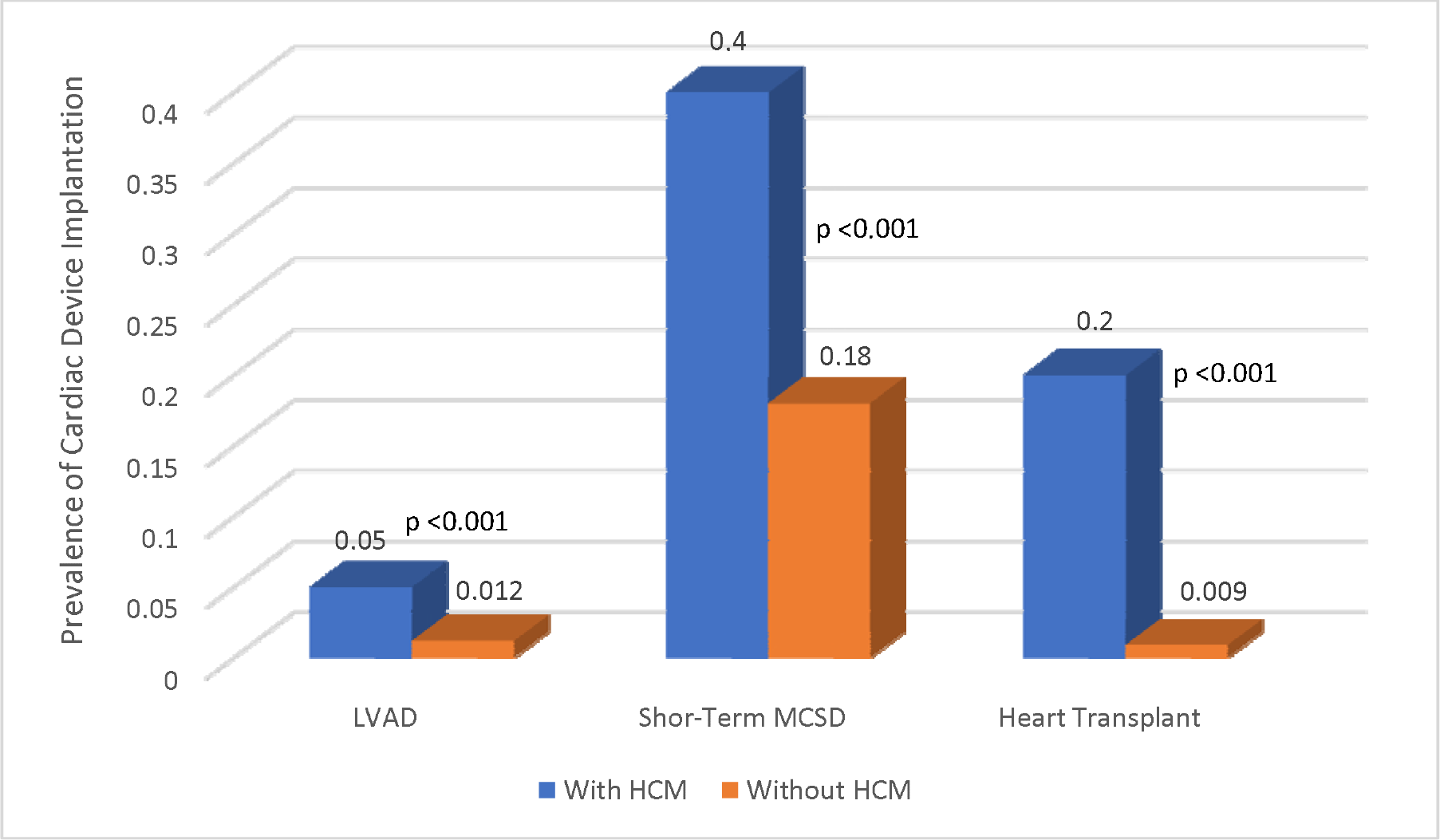
Prevalence of Mechanical Circulatory Support Devices and Heart Transplantation with and without Hypertrophic Cardiomyopathy. Patients with underlying hypertrophic cardiomyopathy (HCM) have higher left ventricular assist device (LVAD), short-term MCSD (IABP, Impella, ECMO) and heart transplantation (HT) rates than those without HCM.

**Figure 2:**
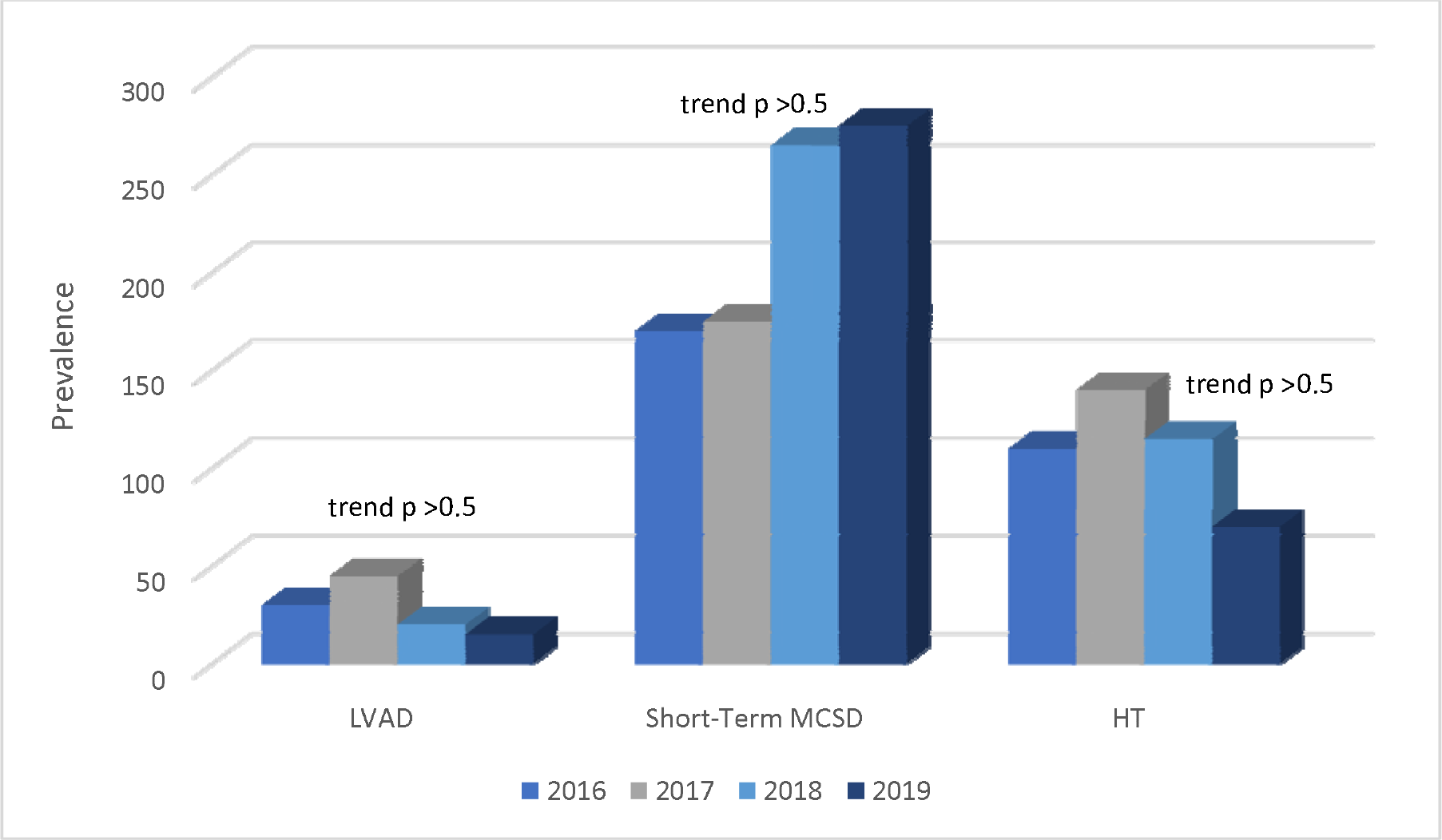
Temporal trends in Mechanical Circulatory Support Devices and Heart Transplantation in Hypertrophic Cardiomyopathy. Left ventricular assist device (LVAD), short-term MCSD (IABP, Impella, ECMO) and heart transplantation (HT) rates among hypertrophic cardiomyopathy (HCM) patients have remained stable (p>0.05)

**Table 2:** Prevalence of MCSD and HT in patients with and without HCM.

| Modality | HCM Patients (n=222945) |  | Non-HCM Patients (n=142000000) |  | p value |
| --- | --- | --- | --- | --- | --- |
|  | Number of Patients (n) | Proportion (%) | Number of Patients (n) | Proportion (%) |  |
| **LVAD** | 110 | 0.05 | 17015 | 0.012 | <0.001 |
| **Short-Term MCSD** | 885 | 0.4 | 253285 | 0.18 | <0.001 |
| **Heart Transplant** | 435 | 0.2 | 12740 | 0.009 | <0.001 |

HCM: Hypertrophic Cardiomyopathy, LVAD: Left Ventricular Assist Device, MCSD: Mechanical Circulatory Support Devices, HT: Heart Transplantation

### Mortality Outcomes of MCSD and HT in HCM

**Table 3** presents the univariate and multivariate logistic regressions for the effect of underlying HCM on mortality in patients undergoing MCSD and HT. Underlying HCM is associated with increased odds of mortality in patients undergoing LVAD implantation (OR 2.43, 95% CI 1.1-6.1, *p*=0.04), an effect preserved after multivariate adjustment (OR 3.06, 95% CI 1.18-7.93, *p*=0.02). Similarly, HCM is associated with increased odds of mortality in patients undergoing MCSD implantation in both univariate (OR 2.25, 95% CI 1.66-3.04, *p*<0.001) and multivariate logistic regressions (OR 1.8, CI 1.29-2.5, p<0.001). Underlying HCM is not associated with increased mortality in patients hospitalized for HT (OR 1.05, CI 0.42-2.6, *p*=0.9).

**Table 3:** Effect of underlying HCM on in-hospital mortality in MCSD and HT.

| Modality | Effect of Underlying HCM on Mortality in CDI |  |  |  |
| --- | --- | --- | --- | --- |
|  | Univariate Logistic Regression |  | Multivariate Logistic Regression |  |
|  | Odd Ratio (95% CI) | p Value | Adjusted Odds Ratio (95% CI) | p Value |
| **LVAD** | 2.43 (1.10-6.11) | 0.04 | 3.06 (1.18-7.93) | 0.02 |
| **Short-Term MCSD (IABP, Impella, ECMO)** | 2.25 (1.66-3.04) | <0.0001 | 1.8 (1.29-2.50) | <0.0001 |
| **Heart Transplant** | 1.05 (0.42-2.60) | 0.09 | - | - |

ECMO: Extracorporeal Membrane Oxygenation, HCM: Hypertrophic Cardiomyopathy, HT: Heart Transplantation, IABP: Intra-Aortic Balloon Pump, LVAD: Left Ventricular Assist Device, MCSD: Mechanical Circulatory Support Devices,

**Figure 3** delineates the temporal trends for adjusted mortality rates for MCSD and HT in patients with and without HCM (**Tables S4**, **S5**). Between 2016 and 2019, patients with HCM experienced a non-significant (*p*_trend_ = 0.09) increase in mortality from 2.5% to 3.5%, whereas their non-HCM counterparts had a significant (*p*_trend_ <0.001) increase in mortality from 1.9% to 2.1%. Patients with HCM hospitalized for MCSD and HT had significantly longer mean hospital lengths of stay as compared to non-HCM patients (26.6 vs 14.4 days, p<0.0001), and higher mean hospitalization charges ($977797 vs $497590, p<0.0001).

**Figure 3:**
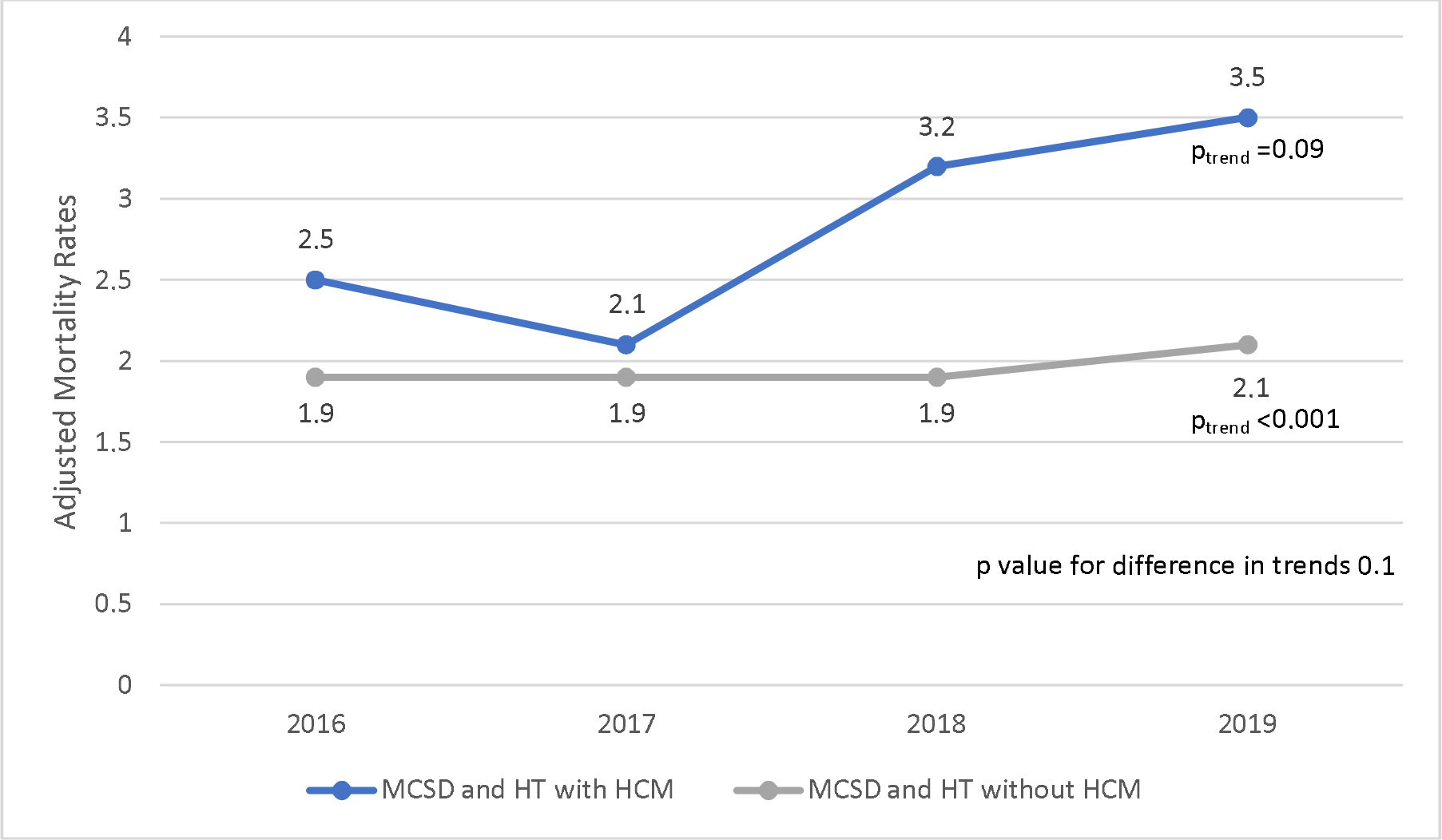
Temporal trends of adjusted mortality rates in Mechanical Circulatory Support Devices and Heart Transplantation with and without Hypertrophic Cardiomyopathy. From 2016 and 2019, hypertrophic cardiomyopathy (HCM) patients had a non-significant (*p*_trend_ = 0.09) increase in mortality, whereas their non-HCM counterparts had a significant (*p*_trend_ <0.001) increase in mortality.

## Discussion

The present study is the first to provide a comprehensive, comparative data analysis of the clinical characteristics and in-hospital outcomes of a HCM population admitted to US hospitals for MCSD and HT. This work builds upon prior HCM investigations of the NIS^8–10^ to expand our understanding of contemporary HCM in-hospital outcomes for MCSD and HT.

We describe the baseline characteristics of patients with HCM receiving MCSD and HT (**Table 1**, **S2**) and trends in prevalence (**Figure 2**, **Table 2**). Notably, the congruent mean age of patients receiving LVADs and heart transplants is in line with the former serving as a bridge therapy to the latter^4–6^. On one hand, the percentage of HCM hospitalizations for MCSD and HT (**Figure 2**) remained stable over the examined time period. This is in agreement with largely unchanged management recommendations over the time period examined (2016-19)^11, 12^. This could also explain the unchanged adjusted, time-dependent in-hospital mortality rates (*p* = 0.09, **Figure 3**), since practice patterns should have accordingly remained relatively unchanged. On the other hand, the prevalence of HCM in the general population increases over the same time frame. This does not necessarily represent a true increase in the epidemiological disease burden, but instead could be explained by an increasing clinical recognition of this condition as many HCM patients can remain undiagnosed throughout life^13^. Regardless, the higher length of stay and hospitalization charges among HCM patients admitted for MCSD and HT compared to their non-HCM counterparts reflects the increased clinical complexity of the condition resulting in a greater healthcare utilization burden. Limited data exists on such healthcare utilization rates in HCM with one study expectedly noting higher HCM costs when a procedure is required, but lacking a comparison group^14^.

Key among the present study’s insights is the increased adjusted in-hospital mortality among HCM patients receiving LVAD or short-term MCSD as compared to their non-HCM counterparts (**Table 2**). The increased mortality risk, persisting even after adjusting for demographic, comorbidity and healthcare setting characteristics, raises the possibility that HCM pathophysiology renders patients a comparatively less optimal candidate for these advanced therapies in cardiogenic shock. As discussed earlier, besides case reports and series^2, 3^, data regarding the in-hospital outcomes related to IABP, Impella or ECMO use in the case of HCM is limited. Sherrid et al^3^ suggested that LVOTO in HCM leads to high LV outflow peak gradient which in association with apical ballooning and wall motion abnormality can develop cardiogenic shock. This systolic dysfunction can be caused by supply-demand ischemia and afterload mismatch^3^. The patients in this case series required vasopressors, short-term MCSD with IABP, Impella, and ECMO. While some of them failed these therapies and needed urgent surgical myomectomy. The patients survived acute shock phase and LV systolic function improved after LVOTO relief^3^. However, there are no known studies comparing the outcomes of MCSD in HCM compared to their non-HCM counter-parts. Hence, the increased risk of in-hospital mortality (adjusted OR 1.8 (1.29-2.50), *p* <0.001, **Table 2**) associated with MCSD use in HCM is a novel finding requiring further investigation to elucidate its underlying etiology.

A prior systematic review found a greater short-term mortality among a pooled cohort of 338 HCM and restrictive cardiomyopathy (RCM) LVAD recipients when compared to ischemic and dilated cardiomyopathy (ICM/DCM) patients^15^. In contrast, a retrospective study of 8 HCM/RCM LVAD recipients showed comparable early (30 days) and late (up to 1 year) mortality outcomes when compared again to their ICM/DCM counterparts^4^. This discrepancy in the literature could reflect differences in critical care practice patterns across US hospitals in patient selection and timing of LVAD placement and/or the difficulty in sufficiently powering a study to detect differences in an otherwise rare clinical occurrence. Regardless, our analysis of 110 LVAD recipients, supports the findings of the former study and additionally localizes the risk to a HCM-specific population, as RCM patients were excluded. The observed greater mortality can be explained by challenges in the applicability of LVADs to HCM pathophysiology, which entails thick LV walls with small associated cavities^15^. Indeed, prior investigations have correlated a smaller LV cavity size with increased mortality post-LVAD implantation in HCM and RCM patients^5, 16^. Further investigations focused on HCM patients are needed to investigate whether the underlying cardiac pathophysiology or any ensuing noncardiac complications^15^ can explain this mortality differential.

A key limitation of the present study is the use of an administrative database relying on billing codes rendering NIS entries susceptible to misclassification error, while the database’s retrospective design precludes causal inferences. Additionally, the NIS lacks data pertaining to laboratory values, pharmacotherapy and imaging studies. Hence, we view this analysis as a tool for hypothesis generation to guide subsequent investigations.

## Conclusion

The present study expands our understanding of in-hospital outcomes and practice patterns of HCM patients receiving MCSD and HT. Key among its insights, this study suggests that LVAD and short-term MCSD use are associated with an increased in-hospital mortality among HCM patients when compared to their non-HCM counterparts. This statistical association is important for clinicians to be aware of when making patient selection decisions for escalation to advanced therapies and should be further investigated in prospective studies.

## Perspectives

### What Is Known?

Hypertrophic cardiomyopathy can uncommonly cause left ventricular outflow tract obstruction and apical ballooning which can present as cardiogenic shock requiring mechanical circulatory support devices

### What Is New?

This study highlights that underlying hypertrophic cardiomyopathy is associated with in-patient hospital mortality, hospitalization charges and length of stay in patients receiving left ventricular assist devices and short term mechanical circulatory support devices including intra-aortic balloon pump, Impella, and extracorporeal membrane oxygenation.

### What Is Next?

Prospective studies are required to further explore the outcomes of hypertrophic cardiomyopathy in mechanical circulatory support devices and establish management guidelines.

## Data Availability

National Inpatient Sample

https://hcup-us.ahrq.gov/nisoverview.jsp

## Abbreviations

ECMO: Extracorporeal Membrane Oxygenation
HCM: Hypertrophic Cardiomyopathy
HT: Heart Transplantation
IABP: Intra-Aortic Balloon Pump
LV: Left Ventricle
LVAD: Left Ventricular Assist Device
LVOT: Left Ventricular Outflow Tract Obstruction
MCSD: Mechanical Circulatory Support Devices

